# Score for Emergency Risk Prediction (SERP): An Interpretable Machine Learning AutoScore–Derived Triage Tool for Predicting Mortality after Emergency Admissions

**DOI:** 10.1101/2021.02.09.21251397

**Authors:** Feng Xie, Marcus Eng Hock Ong, Johannes Nathaniel Min Hui Liew, Kenneth Boon Kiat Tan, Andrew Fu Wah Ho, Gayathri Devi Nadarajan, Lian Leng Low, Yu Heng Kwan, Benjamin Alan Goldstein, David Bruce Matchar, Bibhas Chakraborty, Nan Liu

## Abstract

**Importance:** Triage in the emergency department (ED) for admission and appropriate level of hospital care is a complex clinical judgment based on the tacit understanding of the patient’s likely acute course, availability of medical resources, and local practices. While a scoring tool could be valuable in triage, currently available tools have demonstrated limitations.

**Objective:** To develop a tool based on a parsimonious list of predictors available early at ED triage, to provide a simple, early, and accurate estimate of short-term mortality risk, the Score for Emergency Risk Prediction (SERP), and evaluate its predictive accuracy relative to published tools.

**Design, Setting, and Participants:** We performed a single-site, retrospective study for all emergency department (ED) patients between January 2009 and December 2016 admitted in a tertiary hospital in Singapore. SERP was derived using the machine learning framework for developing predictive models, AutoScore, based on six variables easily available early in the ED care process. Using internal validation, the SERP was compared to the current triage system, Patient Acuity Category Scale (PACS), Modified Early Warning Score (MEWS), National Early Warning Score (NEWS), Cardiac Arrest Risk Triage (CART), and Charlson Comorbidity Index (CCI) in predicting both primary and secondary outcomes in the study.

**Main Outcomes and Measures:** The primary outcome of interest was 30-day mortality. Secondary outcomes include 2-day mortality, inpatient mortality, 30-day post-discharge mortality, and 1-year mortality. The SERP’s predictive power was measured using the area under the curve (AUC) in the receiver operating characteristic (ROC) analysis. Sensitivity, specificity, positive predictive value (PPV), and negative predictive value (NPV) were calculated under the optimal threshold, defined as the point nearest to the upper-left corner of the ROC curve.

**Results:** We included 224,666 ED episodes in the model training cohort, 56,167 episodes in the validation cohort, and 42,676 episodes in the testing cohort. 18,797 (5.8%) of them died in 30 days after their ED visits. Evaluated on the testing set, SERP outperformed several benchmark scores in predicting 30-day mortality and other mortality-related outcomes. Under cut-off score of 27, SERP achieved a sensitivity of 72.6% (95% confidence interval [CI]: 70.7-74.3%), a specificity of 77.8% (95% CI: 77.5-78.2), a positive predictive value of 15.8% (15.4-16.2%) and a negative predictive value of 98% (97.9-98.1%).

**Conclusions:** SERP showed better prediction performance than existing triage scores while maintaining easy implementation and ease of ascertainment at the ED. It has the potential to be widely applied and validated in different circumstances and healthcare settings.

**Key points:** *Question:* How does a tool for predicting hospital outcomes based on a machine learning-based automatic clinical score generator, AutoScore, perform in a cohort of individuals admitted to hospital from the emergency department (ED) compared to other published clinical tools?

*Findings:* The new tool, the Score for Emergency Risk Prediction (SERP), is parsimonious and point-based. SERP was more accurate in identifying patients who died during short or long-term care, compared with other point-based clinical tools.

*Meaning:* SERP, a tool based on AutoScore is promising for triaging patients admitted from the ED according to mortality risk.

## Introduction

Triage in the emergency department (ED) for admission and appropriate level of hospital care is a complex clinical judgment based on the tacit understanding of the patient’s likely acute course, availability of medical resources, and local practices^1,2^. Besides triage categories, early warning scores are also used to identify patients at risk of having adverse events. One such example is the Cardiac Risk Assessment Triage (CART) score ^3^, which calculates a score based on a patient’s vital signs, indicating their risk for cardiac arrest, subsequent transfer to the ICU, and mortality. When a patient presents with suspected cardiac chest pain, such a score has the potential to guide further evaluation and treatment, potentially resulting in fewer adverse events and improved patient outcomes ^4^.

To date, there have been few studies published of predictors of short-term mortality of the general ED population, using the limited data available at the point of triage. Most ED-specific scores are targeted towards specific conditions, such as the quick Sepsis-related Organ Failure Assessment (qSOFA) for infection and sepsis^5,6^, CART for cardiac conditions, or PREDICT for the elderly^7,8^. Two general-purpose scores have been adapted for the ED, such as the Modified Early Warning Score (MEWS) and Acute Physiology and Chronic Health Evaluation (APACHE) II score. However, the MEWS has only moderate predictive capabilities, with an area under the receiver operating characteristic curve (AUC) of 0.71 ^9^, and APACHE II requires laboratory variables not available at the point of triage ^10^. To be useful in the fast-paced ED environment with only limited information, a scoring tool needs to be both accurate and simple.

To address the need for a risk tool appropriate to the ED workflow, we developed the Score for Emergency Risk Prediction (SERP) using a general-purpose machine learning framework we previously described, AutoScore. For this effort, we included all patients who were registered in the ED of a major tertiary hospital in Singapore from January 2009 to December 2016. The primary outcome to be predicted was 30-day mortality. The predictions were based on six data elements easily attainable at triage, including vital signs and comorbidities. The resulting tool was compared in a test set to the current triage system used in Singapore, the Patient Acuity Category Scale (PACS) ^11^, as well as three published early warning or triage scores.

## Methods

### Study design and setting

We performed a retrospective cohort study of patients seen in the ED of Singapore General Hospital (SGH). Singapore is a city-state in Southeast Asia, facing a rapidly aging society ^12^; currently, about 1 in 5 Singaporeans are aged 60 or above ^13^. SGH is the largest and oldest public tertiary hospital in Singapore. The SGH ED receives over 120,000 visits and has 36,000 in-patient admissions annually. Electronic Health Record (EHR) data were obtained from Singapore Health Services and employed in this study. This study was approved by Singapore Health Services’ Centralized Institutional Review Board, and a waiver of consent was granted for EHR data collection.

### Study population

All patients visiting SGH ED from January 1, 2009 until December 31, 2016, who were subsequently admitted were included. We denote these included episodes as emergency admissions. Patients below 21 years old or who died in the ED were excluded. We also excluded foreign patients who may not have complete medical records. Admission episodes from January 1, 2009 to December 31, 2015 were randomly split into 2 non-overlapping cohorts: a training cohort (80%) and a validation cohort (20%). The admission episodes dated between January 1 and December 31, 2016 were assigned to the testing cohort. This sequential testing design was chosen to be more consistent with real future application scenarios as well as to test whether the population shift would influence the model’s performance

### Outcome

The primary outcome used to develop and test the tool was 30-day mortality, defined as deaths within 30 days after the date of emergency admission. Secondary outcomes included inpatient mortality, defined as deaths in the hospital; 2-day mortality, defined as deaths within 48 hours after the time of admission; 30-day post-discharge mortality, defined as deaths within 30 days after the date of hospital discharge; 1-year mortality, defined as deaths within 365 days after the date of emergency admission. Death records were obtained from the national death registry and were matched to specific patients in the EHR.

### Data collection and candidate variables

We extracted data from the hospital’s EHR, through the SingHealth Electronic Health lntelligence System (eHints). Patients’ details were de-identified, complying with HIPAA regulations. Comorbidities were obtained from the hospital diagnosis and discharge records in the preceding five years before patients’ index emergency admissions. They were extracted from the International Classification of Diseases (ICD) codes (ICD-9/ICD-10) ^14^, which is a globally used diagnostic tool for epidemiology and clinical purposes. We preselected candidate variables that are available in the ED before hospital admission to ensure SERP is clinically useful for early risk stratification of patients at the ED. Candidate variables included demographics, administrative variables, medical history in the preceding year, clinical vital signs, and comorbidities. The list of candidate variables was shown in the web-only appendices (eTable 1). Comorbidities variables were defined according to the Charlson Comorbidity Index (CCI). We used the algorithms developed and updated by Quan and colleagues ^15^ for the linkage between CCI and ICD codes.

**Table 1:** Basic characteristics of the study cohort

|  | Training cohort | Validation Cohort | Testing Cohort |
| --- | --- | --- | --- |
| *n* | 224666 | 56167 | 42676 |
| **Demographics** |  |  |  |
| Age (yr), mean (SD) | 63.60 (16.90) | 63.58 (16.87) | 64.85 (16.80) |
| Gender, *n* (%) |  |  |  |
| Male | 111240 (49.5) | 27740 (49.4) | 21120 (49.5) |
| Race, *n* (%) |  |  |  |
| Chinese | 167004 (74.3) | 41765 (74.4) | 31441 (73.7) |
| Indian | 22403 (10.0) | 5592 (10.0) | 4440 (10.4) |
| Malay | 29040 (12.9) | 7213 (12.8) | 5465 (12.8) |
| Others | 6219 (2.8) | 1597 (2.8) | 1330 (3.1) |
| PACS triage categories, *n* (%) |  |  |  |
| P1 | 39548 (17.6) | 9823 (17.5) | 9913 (23.2) |
| P2 | 128644 (57.3) | 32058 (57.1) | 22885 (53.6) |
| P3 and P4 | 56474 (25.1) | 14286 (25.4) | 9878 (23.1) |
| Shift time, *n* (%) |  |  |  |
| 08:00 to 16:00 | 113758 (50.6) | 28461 (50.7) | 21870 (51.2) |
| 16:00 to 24:00 | 84503 (37.6) | 21050 (37.5) | 15907 (37.3) |
| 24:00 to 8:00 | 26405 (11.8) | 6656 (11.9) | 4899 (11.5) |
| Day of week, *n* (%) |  |  |  |
| Friday | 31553 (14.0) | 7893 (14.1) | 5839 (13.7) |
| Monday | 37703 (16.8) | 9581 (17.1) | 7139 (16.7) |
| Weekend | 57785 (25.7) | 14283 (25.4) | 10901 (25.5) |
| Midweek | 97625 (43.5) | 24410 (43.5) | 18797 (44.0) |
| **Vital signs** |  |  |  |
| Pulse (beat/min), mean (SD) | 81.57 (16.41) | 81.62 (16.37) | 85.95 (18.36) |
| Respiration (breath/min), mean (SD) | 17.80 (1.57) | 17.80 (1.59) | 18.23 (2.04) |
| SpO<sub>2</sub> (%), mean (SD) | 98.12 (2.84) | 98.12 (2.70) | 97.34 (4.18) |
| Diastolic BP (mmHg), mean (SD) | 70.99 (13.23) | 71.01 (13.20) | 72.36 (13.95) |
| Systolic BP (mmHg), mean (SD) | 133.77 (24.49) | 133.80 (24.58) | 137.73 (27.87) |
| **Comorbidities** |  |  |  |
| Myocardial infarction, *n* (%) | 14927 (6.6) | 3801 (6.8) | 2841 (6.7) |
| Congestive heart failure, *n* (%) | 28511 (12.7) | 7136 (12.7) | 4897 (11.5) |
| Peripheral vascular disease, *n* (%) | 14531 (6.5) | 3539 (6.3) | 2541 (6.0) |
| Stroke, *n* (%) | 32993 (14.7) | 8062 (14.4) | 5062 (11.9) |
| Dementia, *n* (%) | 6901 (3.1) | 1699 (3.0) | 1515 (3.6) |
| Chronic pulmonary disease, *n* (%) | 24275 (10.8) | 6138 (10.9) | 3912 (9.2) |
| Rheumatoid disease, *n* (%) | 3341 (1.5) | 881 (1.6) | 615 (1.4) |
| Peptic ulcer disease, *n* (%) | 9879 (4.4) | 2505 (4.5) | 1362 (3.2) |
| Diabetes, *n* (%) |  |  |  |
| Nil | 145889 (64.9) | 36457 (64.9) | 27204 (63.7) |
| Diabetes without chronic complications | 24268 (10.8) | 6064 (10.8) | 1247 (2.9) |
| Diabetes with complications | 54509 (24.3) | 13646 (24.3) | 14225 (33.3) |
| Hemiplegia or paraplegia, *n* (%) | 14545 (6.5) | 3609 (6.4) | 1880 (4.4) |
| Renal disease, *n* (%) | 49884 (22.2) | 12483 (22.2) | 10377 (24.3) |
| Cancer, *n* (%) |  |  |  |
| Nil | 185121 (82.4) | 46251 (82.3) | 35374 (82.9) |
| Local tumor, leukemia, lymphoma | 20838 (9.3) | 5136 (9.1) | 3613 (8.5) |
| Metastatic solid tumour | 18707 (8.3) | 4780 (8.5) | 3689 (8.6) |
| Liver disease, *n* (%) |  |  |  |
| Nil | 209865 (93.4) | 52562 (93.6) | 39704 (93.0) |
| Mild liver disease | 11112 (4.9) | 2676 (4.8) | 2156 (5.1) |
| Severe liver disease | 3689 (1.6) | 929 (1.7) | 816 (1.9) |
| **Medical history** |  |  |  |
| Count of emergency admissions last year, mean (SD) | 1.05 (2.35) | 1.05 (2.35) | 1.12 (2.51) |
| Count of surgeries last year, mean (SD) | 0.20 (0.72) | 0.20 (0.72) | 0.28 (0.94) |
| Count of ICU admission last year, mean (SD) | 0.03 (0.26) | 0.02 (0.26) | 0.03 (0.29) |
| Count of HD admissions last year, mean (SD) | 0.10 (0.51) | 0.10 (0.51) | 0.08 (0.45) |
| **Mortality-related outcomes** |  |  |  |
| Inpatient Mortality, *n* (%) | 8616 (3.8) | 2151 (3.8) | 1515 (3.6) |
| 2-day mortality, *n* (%) | 1801 (0.8) | 449 (0.8) | 295 (0.7) |
| 30-day post-discharge mortality, *n* (%) | 7742 (3.6) | 1905 (3.5) | 1343 (3.3) |
| 1-year mortality, *n* (%) | 45013 (20.0) | 11015 (19.6) | 8185 (19.2) |
| 30-day mortality, *n* (%) | 13244 (5.9) | 3285 (5.8) | 2310 (5.4) |

ED=Emergency Department, SD= Standard Deviation, ED=Emergency Department, BP=Blood Pressure, SpO<sub>2</sub>= Blood Oxygen Saturation, LOS= Length of stay

### Statistical analysis

The data were analyzed using R 3.5.3 (R Foundation, Vienna, Austria). Baseline characteristics of the study population were analyzed on all three cohorts to confirm their similarity. In the descriptive summaries, frequencies and percentages were reported for categorical variables. For continuous variables, means and standard deviations (SDs) were reported. During the analysis, value for vital signs would be considered as an outlier and set to missing if it was beyond the plausible physiological ranges based on domain knowledge, such as any value of vital signs below zero, heart rate above 300, respiration rate above 50, systolic blood pressure above 300, diastolic blood pressure above 180 or SpO_2_ above 100. Then, all missing values were imputed using the median value of the training cohort.

We implemented the AutoScore ^16^, a machine learning-based clinical score generation algorithm to derive the SERP scoring model. AutoScore combines both machine learning and logistic regression, integrates multiple modules of data manipulation, and automates the development of parsimonious sparse-score risk models for predefined outcomes. Also, it enables users to build transparent and straightforward clinical scores quickly and seamlessly, which can be easily implemented and validated in clinical practice. The training cohort was used for the generation of the tentative SERP models using AutoScore main flow. The validation cohort was utilized to evaluate multiple candidate SERP models for parameter tuning and model selection. Then, we calculated the performance metrics of the final SERP model based on the testing cohort. All implementation details and methodology descriptions were shown in the web-only Appendices. We used the primary outcomes for model derivation and applied all outcomes for model testing. The implementation details and mythological descriptions were attached in the web-only appendices (eFigure1 and eTextbox).

After model derivation, the predictive performance of the final SERP model was reported based on the testing cohort, and bootstrapped samples were applied to calculate 95% confidence intervals (CIs). Each of the SERP breakdowns should be allocated a score reflecting the magnitude of disturbance to each variable. The individual scores should then be summed up to derive the aggregate SERP score for risk stratification of outcomes. The predictive power of SERP was measured using the area under the curve (AUC) in the receiver operating characteristic (ROC) analysis. Sensitivity, specificity, positive predictive value (PPV), and negative predictive value (NPV) were calculated under the optimal threshold, defined as the point nearest to the upper-left corner of the ROC curve. The metrics calculated under different thresholds were also compared to evaluate predictive performance. By using the same testing cohort, we compared the SERP with the PACS, Modified Early Warning Score (MEWS)^17^, National Early Warning Score (NEWS)^18^, Cardiac Arrest Risk Triage (CART)^19^, and Charlson Comorbidity Index (CCI)^20^ in predicting both primary and secondary outcomes in the study.

## Results

### Baseline characteristics of the study cohort

Between January 2009 and December 2015, a total of 280,833 individual admission episodes were included (224,666 in the training cohort and 56,167 in the validation cohort). Besides, 42,676 admission episodes in the year 2016 were included in the testing cohort (Figure 1). The mean age of the main training cohort was 63.6 (SD = 16.9) and 49.5% were male (n=111,240). The ethnic compositions were similar to the population norm (74.3% for Chinese, 12.9% for Malay, 10.0% for Indian, and 2.8% for others). 17.6% (n = 39548) of episodes were triaged as PACS 1 and 57.3% (n = 128,644) of episodes were triaged as PACS 2. The mean ED boarding time was 4.72 (SD = 3.99) hours, and ED consultation waiting time was 0.8 (SD = 0.76) hours. Table 1 shows the populations in both training and validation cohorts, which were similar in terms of age, gender, ethnic compositions, and other characteristics. Compared with the training and validation cohorts, however, patients in the testing cohort were slightly older, had a higher risk for triage to PACS 1, with more people having the comorbidities of myocardial infarction, diabetes, renal diseases. The patients in the testing cohort also had marginally lower mortality rates while having higher numbers of emergency admissions or surgeries in the past year. This likely reflects the population shift and improvements in healthcare over time.

**Figure 1:** Flow of the cohort formation.

### Selected variables and SERP score

AutoScore was used to select the most discriminative variables from all 26 candidate variables (eTable 1). A parsimony plot (i.e., model performance vs. complexity) based on the validation set was used for determining the choice of variables (eFigure 2). We chose six variables as the parsimonious choice as it achieved a good balance in the parsimony plot. These six variables were: age, heart rate, respiration rate, diastolic blood pressure, systolic blood pressure, and history of cancer (including local tumor, leukemia, lymphoma, and metastatic solid tumor). Those selected variables would highlight the importance of vital signs in risk-triaging patients in emergency settings. As seen from eFigure 2, when more variables were added to the scoring model, the performance was not markedly improved.

The SERP, a six-variable scoring model is tabulated in Figure 2. The final score summed up from six variables ranged from 0 to 60. We used the testing cohort to evaluate the performance of the SERP score. eFigure 3 depicts the distribution of episodes at different score intervals, which were near-normal distribution. Most patients had a risk score between 16 and 24, and very few patients had scores under nine or above 40. As seen from eFigure 4, the observed mortality rate increased as our risk scores grew on the testing cohort. The observed mortality rate was about 5% for the score of 27, while the mortality rate was over 20% for the score of above 36. In terms of different breakdowns of the SERP, when age was lower than 30, its corresponding risk (quantified as points) was the lowest; when it was higher than 80, the risk was the highest. Likewise, when a reported diastolic BP was between 50 and 94, the corresponding risk was the lowest, and when it was lower than 49, the risk was the highest. Also, some variables have larger score values, elucidating more significant contributions to the score, such as age, heart rate, and comorbidity of different types of cancers.

**Figure 2:** Six-variable Score for Emergency Risk Prediction (SERP)

**Figure 3:**
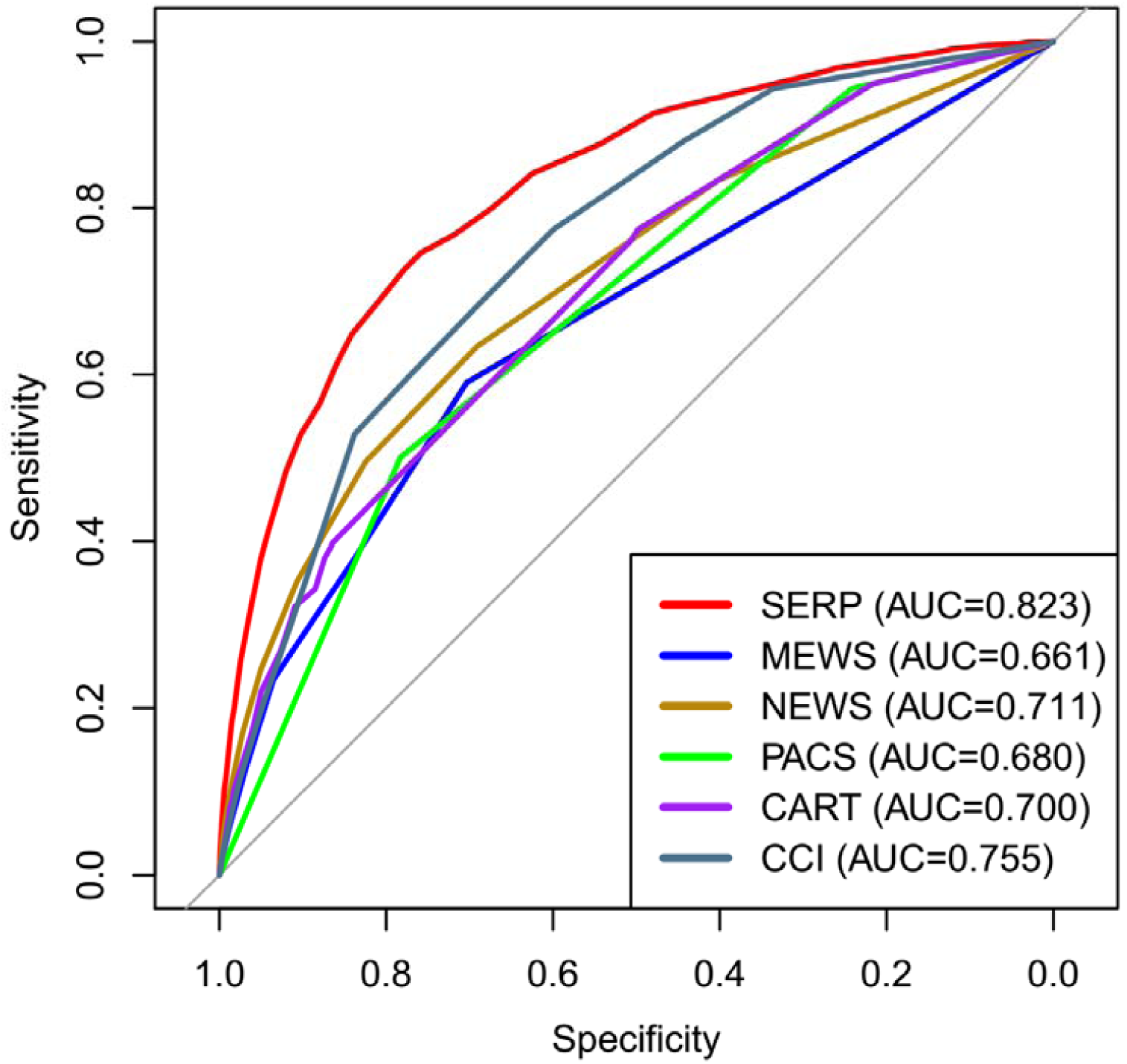
Receiver operating characteristic curves of SERP and other benchmark models. AUC, area under the curve; SERP, Score for Emergency Risk Prediction; MEWS, Modified Early Warning Score; PACS, Singapore local-based Patient Acuity Category Scale; NEWS, National Early Warning Score; CART, Cardiac Arrest Risk Triage; CCI, Charlson Comorbidity Index.

### Performance evaluation

The performance of the SERP score and other clinical scores, as assessed by ROC analysis on the testing cohort, are reported in Table 2 and Figure 3.

**Table 2:** Comparison of AUC values achieved by different triage scores on the testing set.

|  | **30-day**<br>**Mortality** | **2-day**<br>**Mortality** | **Inpatient**<br>**Mortality** | **30day-Post-**<br>**discharge**<br>**Mortality** | **1-year**<br>**Mortality** |
| --- | --- | --- | --- | --- | --- |
| **SERP** | 0.823 (0.814-0.832) | 0.821 (0.796-0.847) | 0.810 (0.799-0.821) | 0.814 (0.802-0.826) | 0.785 (0.78-0.791) |
| **PACS** | 0.680 (0.670-0.69) | 0.796 (0.775-0.817) | 0.703 (0.691-0.715) | 0.381 (0.368-0.393) | 0.390 (0.384-0.396) |
| **MEWS** | 0.663 (0.652-0.674) | 0.763 (0.734-0.792) | 0.680 (0.667-0.694) | 0.613 (0.598-0.627) | 0.583 (0.577-0.589) |
| **NEWS** | 0.711 (0.700-0.723) | 0.803 (0.774-0.832) | 0.734 (0.72-0.747) | 0.655 (0.64-0.67) | 0.623 (0.616-0.629) |
| **CART** | 0.700 (0.689-0.711) | 0.779 (0.751-0.807) | 0.704 (0.691-0.717) | 0.665 (0.651-0.679) | 0.652 (0.646-0.658) |
| **CCI** | 0.755 (0.746-0.765) | 0.687 (0.659-0.715) | 0.743 (0.731-0.755) | 0.777 (0.765-0.789) | 0.786 (0.781-0.792) |

AUC, area under the curve; SERP, Score for Emergency Risk Prediction; PACS, Patient Acuity Category Scale; MEWS, Modified Early Warning Score; NEWS, National Early Warning Score; CART, Cardiac Arrest Risk Triage; CCI, Charlson Comorbidity Index.

SERP showed promising discriminatory capability in predicting all mortality-related outcomes. It achieved an AUC of 0.823 (95% CI: 0.814-0.832) for 30-day mortality, an AUC of 0.821 (95% CI: 0.796-0.847) for 2-day mortality, an AUC of 0.81 (95% CI: 0.799-0.821) for inpatient mortality, an AUC of 0.814 (95% CI: 0.802-0.826) for 30-day-post-discharge mortality, and an AUC of 0.785 (95% CI: 0.78-0.791) for 1-year mortality. In comparison, none of the other clinical scores could achieve an AUC of more than 0.8 in any mortality-related outcome. eTable 3 presents varying thresholds of predicted risk based on the SERP, the proportion of patients stratified for 30-day mortality, and corresponding sensitivity, specificity, positive and negative predictive values. eTable 2 also presents varying thresholds of predicted risk based on other comparators, including MEWS, NEWS, CART, and CCI. Based on ROC analysis, the optimal cut-off of the SERP model is 26, which is located nearest to the upper-left corner of the ROC curves. The calibration curve of the SERP model was shown in eFigure 5.

## Discussion

In this study, we developed SERP, a parsimonious and point-based scoring tool for triaging patients at the ED. SERP is more accurate in identifying patients who died during short or long-term care, compared with other point-based clinical tools (i.e., PACS, NEWS, MEWS, CART, and CCI). We previously developed a model for inpatient mortality using variables consisting of basic demographic, administrative and clinical information acquired in ED^21^. Despite the model showing good discriminative performance, the need to use a computer with the 19 variables and limited its applicability and interpretability. Instead, SERP is an additive, point-based triage tool, making it simple, quick to calculate, transparent, and interpretable. SERP has the advantage of easy implementation and interpretation and thus could be widely utilized and validated in different circumstances.

This study has several strengths. Machine learning-based variable selection by AutoScore^16^ can efficiently filter out redundant information to achieve a sparse solution. Sanchez-Pinto et al.^22^ also suggested that variable selection plays an essential role in reducing the complexity of prediction models without compromising their accuracy, especially when facing a large number of candidate features extracted from EHRs^23^. Likewise, Liu et al.^24^ demonstrated that more predictors did not necessarily lead to better prediction of adverse cardiac events. The second strength of SERP is the size of the dataset that was used to derive this score. This is one of the largest datasets used to generate a point-based triage model, with a cohort of over 300,000 emergency admissions over eight years, obtained from a large tertiary hospital and representing the population norm. Third, the SERP score consistently performed well in the test cohort, even with changes in patient characteristics, outcome prevalence, and clinical practices amidst the continuously evolving^25^ clinical environment.

There are several possible reasons for SERP to excel in predicting both short and long-term mortality. The SERP score includes comorbidities, the importance of which has been demonstrated in several studies. For example, in a study^26^ by Chu and colleagues, patient comorbidity contributed to both short and long-term mortality. Fortin et al.^27^ also indicated that failure to consider comorbidity might result in biased analyses, possibly due to confounding differences in health status among populations. Furthermore, we included age as a predictor in the SERP score, the importance of which was also highlighted in many studies ^28,29^. Vilpert et al.^28^ have previously shown that the ED is affected by an aging population, with the elderly more susceptible to episodes of acute age-related illness or acute exacerbations of chronic illnesses. This is also reflected in a study by Parker et al.^30^, where increasing age was the strongest predictor for hospital admission besides triage acuity.

Researchers have created many point-based triage tools for the prediction of short or long-term mortality. However, the majority of internationally recognized scores are either not derived in the ED setting, such as APACHE II ^10^ and MEWS ^9^, or are applied only to specific subsets of the ED population, such as the qSOFA^5^ score for sepsis and CART ^3^ score for cardiac conditions. As none of these international mortality prediction scores were explicitly built for mortality prediction in the general ED population, they understandably demonstrate limited predictive capabilities. In contrast, our SERP score can be widely and rapidly utilized in most ED populations in hospitals worldwide, requiring only two simple history questions (age, history of cancer) and three vital sign measurements (pulse rate, blood pressure, respiration rate). Our score can be easily calculated by trained medical assistants or integrated into an existing hospital EHR, allowing for the quick determination of a patient’s mortality risk without adversely affecting ED workloads. This is important in the fast-paced ED environment as well as in heterogenous ED systems around the world, where generalists rather than emergency medicine specialists sometimes run it.

The SERP score provides a concrete measure during the ED triage to assess a patient’s mortality risk. While clinicians are generally able to ascertain the severity of a patient’s acute condition and the threat to life, their decisions are often subjective and depend on an individual’s experience and knowledge. In a study^31^ of elderly patients, while physicians could predict the 30-day mortality of such patients with an odds ratio of 2.4 during the consultation, they missed four out of every five deaths, with a sensitivity of only 20%. In another study^32^, researchers attempted to adopt fast and objective physiological biomarkers to stratify chest pain patients at ED triage according to the risk of 30-day mortality and other adverse cardiac events. Such studies highlight the role of data-driven, objective clinical decision tools to help clinicians rethink and reassess such patients, minimizing the likelihood of such patients falling through the cracks. Besides, various triage scores, like the Emergency Severity Index (ESI)^33^, also includes a highly variable parameter that is affected by the experience of the nurse. In comparison, SERP may potentially bypass this, better-enabling nurses in the rapid triaging of patients.

Given the relatively good prediction performance and advantages of SERP, prospective studies will be designed to further validate its performance for implementation into real-life practice. Also, given the strengths of SERP as a simple yet understandable scoring tool, further assessments must be carried out to evaluate the more intangible aspects of score implementation^34,35^. Such measures would include a determination of SERP’s long-term sustainability, overall cost-effectiveness, and most importantly, physician perceived acceptability and score take-up rates. We believe that these assessments will likely lend credence to SERP as an effective and accurate tool for decision making within the ED.

### Limitations

This study has several limitations. First, the dataset used in this study was based on EHR data of routinely collected variables. Thus, some variables such as comorbidities and prior hospitalizations might not be easily extracted from different EHR systems during external validation of the SERP. Second, as this was a single-center study at a tertiary hospital, the performance of SERP may vary in other settings. Last, the ED cohort only accounted for ED admissions, which might miss the ED visits for individuals not subsequently admitted to the hospital, which might influence generalizability based on the threshold for admission decision in one hospital vs. another. Nevertheless, given our tool’s purpose as an adjunct to clinical acumen during the consultation, we believe that such a mortality risk stratification tool would conceivably be used when a physician is planning to admit a patient and is considering the level of service that might be appropriate for that individual.

## Conclusions

We derived SERP, a parsimonious and point-based scoring tool for triaging patients at the ED. In comparison, SERP performs better than existing triage scores and has the advantage of easy implementation, ease of ascertainment at ED presentation, with the potential to be widely utilized and validated in different circumstances and healthcare settings. Following the clinical application of SERP in ED triage, more tailored scores are potential to be derived in different clinical areas through the data-driven approach by AutoScore in the future.

## Data Availability

Details of the variables and derived predictive model
are available from the corresponding author.

## Author contributions

FX, MEHO, BC, and NL contributed to the study conception and design. FX and JNMHL performed the analyses. The first draft of the manuscript was written by FX, JNMHL, and NL. All authors contributed to the evaluation of the methods, interpretation of the results, and revision of the manuscript. All authors have read and approved the final manuscript for submission. NL takes responsibility for the paper as a whole.

## Funding and support

This research received funding from Duke-NUS Medical School and the Estate of Tan Sri Khoo Teck Puat under the Khoo Pilot Award (Collaborative). This study was also supported by the Singapore National Medical Research Council under the PULSES Center Grant.

## Notes

### Competing Interest Statement

The authors have declared no competing interest.

### Author Declarations

This study was approved by Singapore Health Services' Centralized Institutional Review Board, and a waiver of consent was granted for EHR data collection.

